# Genetic Profiles in Pharmacogenes Indicate Personalized Drug Therapy for COVID-19

**DOI:** 10.1101/2020.03.23.20041350

**Authors:** Lei-Yun Wang, Jia-Jia Cui, Qian-Ying OuYang, Yan Zhan, Yi-Min Wang, Xiang-Yang Xu, Cheng-Xian Guo, Ji-Ye Yin

## Abstract

**Background:** The coronavirus disease 2019 (COVID-19) has become a global pandemic currently. Many drugs showed potential for COVID-19 therapy. However, genetic factors which can lead to different drug efficiency and toxicity among populations are still undisclosed in COVID-19 therapy.

**Methods:** We selected 67 potential drugs for COVID-19 therapy (DCTs) from clinical guideline and clinical trials databases. 313 pharmaco-genes related to these therapeutic drugs were included. Variation information in 125,748 exomes were collected for racial differences analyses. The expression level of pharmaco-genes in single cell resolution was evaluated from single-cell RNA sequencing (scRNA-seq) data of 17 healthy adults.

**Results:** Pharmacogenes, including CYP3A4, ABCB1, SLCO1B1, ALB, CYP3A5, were involved in the process of more than multi DCTs. 224 potential drug-drug interactions (DDIs) of DCTs were predicted, while 112 of them have been reported. Racial discrepancy of common nonsynonymous mutations was found in pharmacogenes including: VDR, ITPA, G6PD, CYP3A4 and ABCB1 which related to DCTs including ribavirin, α-interferon, chloroquine and lopinavir. Moreover, ACE2, the target of 2019-nCoV, was only found in parts of lung cells, which makes drugs like chloroquine that prevent virus binding to ACE2 more specific than other targeted drugs such as camostat mesylate.

**Conclusions:** At least 17 drugs for COVID-19 therapy with predictable pharmacogenes should be carefully utilized in risk races which are consisted of more risk allele carriers. At least 29 drugs with potential of DDIs are reported to be affected by other DDIs, they should be replaced by similar drugs without interaction if it is possible. Drugs which specifically targeted to infected cells with ACE2 such as chloroquine are preferred in COVID-19 therapy.

## Introduction

In the end of last year, Wuhan (the capital city of Hubei province in China) reported a new unknown viral pneumonia ^[1]^. Subsequent next generation sequencing identified that it was caused by a novel coronavirus which named as 2019 novel coronavirus (2019-nCoV) ^[2]^. World Health Organization (WHO) named this new disease as coronavirus disease 2019 (COVID-19) and declared a Public Health Emergency of International Concern. Although it is firstly detected in China, epidemiological monitoring indicates that COVID-19 is experiencing quickly outbreak across the world. As of 22 March 2020, more than 315,925 cases of infection and 13,610 cases of death have been confirmed in at least 184 countries.

Clinical characteristics reporting in China revealed that most patients are mild and moderate ^[3, 4]^. However, severe cases progress rapidly to acute respiratory distress syndrome (ARDS), shock, multiple organ failure and even death. The National Health Commission of China released at least seven versions of Guideline of Diagnosis and Treatment of Pneumonitis Caused by COVID-19 ^[5]^. Oxygen therapy, mechanical ventilation and drug therapy are recommended as major treatments, so that several drugs may be utilized simultaneously in such conditions. It is noteworthy that the individual difference of drug treatment is mentioned for a special in the guideline. This promotes the importance of personalized therapy for COVID-19 patients.

It is widely accepted that genetic factor is one of the major contributors to individual or ethnical differences of drug therapeutic efficacy and toxicity ^[6, 7]^. One of the examples is chloroquine, which is listed as an antiviral drug for COVID-19 treatments in China. Currently, at least 15 clinical trials are being conducted to explore its efficacy. However, existed results show that glucose-6-phosphate dehydrogenase (G6PD) deficient patients are at an increased risk of life-threatening severe hemolysis after taking chloroquine ^[8]^. As one of the most common human enzyme defects, genetic variants affecting G6PD activity show remarkably individual and ethnical differences ^[9]^. Thus, it should be widely informed that chloroquine needs precision medicine.

In addition to chloroquine, a number of other drugs are also used in the COVID-19 therapy. With the rapid spread of this disease in the worldwide, these drugs will be used in patients with different ethnic backgrounds. However, the genetic variants potentially affect their effects and safety are not still available. Their distributions in different populations in the world are also not indicated. This may constitute an obstacle for successful treatment and control of this disease. Pharmacogenetics (PGx) investigates the affection of genetic variants on drug effects and safety. In the current study, we provide a PGx landscape of drugs with potential to be used in the COVID-19 treatment. Candidate choices from different drugs based on pharmacogenetic analyses were provided.

## Methods

### Data collection

Drugs and their clinical trial information for COVID-19 treatment were col lected from Guideline of Diagnosis and Treatment of Pneumonitis Caused by COVID-19 (version 7.0) (http://www.nhc.gov.cn/yzygj/s7653p/202003/46c9294a7dfe4cef80dc7f5912eb1989.shtml), Clinical Trails (https://clinicaltrials.gov) and Chine se Clinical Trail Registry (http://www.chictr.org.cn). The pharmacogenes related to all drugs were obtained from DrugBank (https://www.drugbank.ca/) and Phar mGKB (https://www.pharmgkb.org/) databases. Actionable PGx biomarkers were determined by Clinical Pharmacogenetics Implementation Consortium (CPIC: http://cpicpgx.org/guidelines/) and DPWG (http://clivar-dpwg.iri.columbia.edu/) guid elines. Genetic variation data on each of the genes were retrieved from Gnom AD database (http://gnomad.broadinstitute.org/, version: 2.1.1). Finally, single-cel l RNA sequencing (sc-RNA seq) data from two organs (liver (GSE115469, n=5) and lung (GSE130148, n=4)) of 9 healthy adults was download from GEO da tabase (https://www.ncbi.nlm.nih.gov/gds).

### Sc-RNA seq data analysis

The raw count matrix (unique molecular identifier counts per gene per cell) was processed by Seurat v3 ^[12]^. Firstly, low-quality data was filtered out as following: (1) cells expressed less than 10 genes; (2) genes expressed in less than two percent of cells. Secondly, we calculated log_2_(count-per-million + 1) expression, and followed by normalization and standardization. Thirdly, the cells were clustered. In detail, the highly variable genes were identified and cells were then clustered through embedding them into a graph structure in Principal Component Analysis space. The parameter resolution was set as 0.5 to identify only major cell types. The clustered cells were then projected onto a two-dimensional space and visualized. Finally, the differentially expressed genes on each cluster were identified. They were used to annotate and merge the cell clusters according to curated known cell markers (http://biocc.hrbmu.edu.cn/CellMarker/index.jsp).

### Genetic variations annotation and drug-gene network construction

All genetic variations were annotated by allele frequency, location and function in different populations using ANNOVAR (version: 2019Oct24) (Table S1). In the current study, populations were divided into eight categories: African, Latino, East Asian, South Asian, Finnish, Non-Finnish European, Ashkenazi Jewish and Other. Based on the location and functions, all mutations were divided into 12 categories: frameshift, non-frameshift, synonymous single nucleotide polymorphism (SNP), nonsynonymous SNP, stop-gain, untranslated regions, intronic and others (including splicing, upstream of a gene, downstream of a gene, stop-loss and ncRNA_exonic mutations). Functional nonsynonymous mutations were predicted by PROVEAN (http://provean.jcvi.org/). The drug-gene network was constructed by Cytoscape software (version:3.7.1) ^[10]^.

## Results

### Candidate drugs and their clinical trials for COVID-19 treatment

In the current study, a total of 67 drugs were collected in our study, including agents being used clinically, undergoing clinical trials, only confirmed in *vitro* experiments or simply with the therapeutic potential. They could be classified into nine categories according to their functions: viral toxicity (38.8%) and their sensitizers (3.0%), inhibiting virus invasion (6.0%), improving lung function (4.5%), maintaining tissue and organ function balance (25.4%), anti-hypoxic (4.5%), anti-septic shock (7.5%), anti-secondary infection (6.0%) and others (4.5%) (Figure 1A). Among them, antiviral drugs were mostly important and account for 47.8% in all drugs (including virus invasion inhibitors, viral toxicity and their sensitizers). As of March 18, 2020, a total of 125 clinical trials have been conducted to study the safety and efficacy of antiviral drugs. And most of these clinical trials are all ongoing and have not been completed. There are six clinical trials involving remdesivir, the earliest closed one involving 308 participants will be completed on April 27, 2020 (NCT04252664). As many as 16 clinical trials related to chloroquine, and two clinical trials for mild and common patients and severe patients will be completed on April 30, 2020 (ChiCTR2000029898, ChiCTR2000029899), each clinical trial includes a total of 100 participants. As for hydroxychloroquine, two of the nine clinical trials have ended on February 29, 2020 (ChiCTR2000029559, ChiCTR2000029740), and the results have not yet been available. In addition, the clinical trials of lopinavir (18) and ritonavir (23) will both be completed as early as March 25, 2020. Information of other categories of drugs can be found in Figure S1.

**Figure 1:**
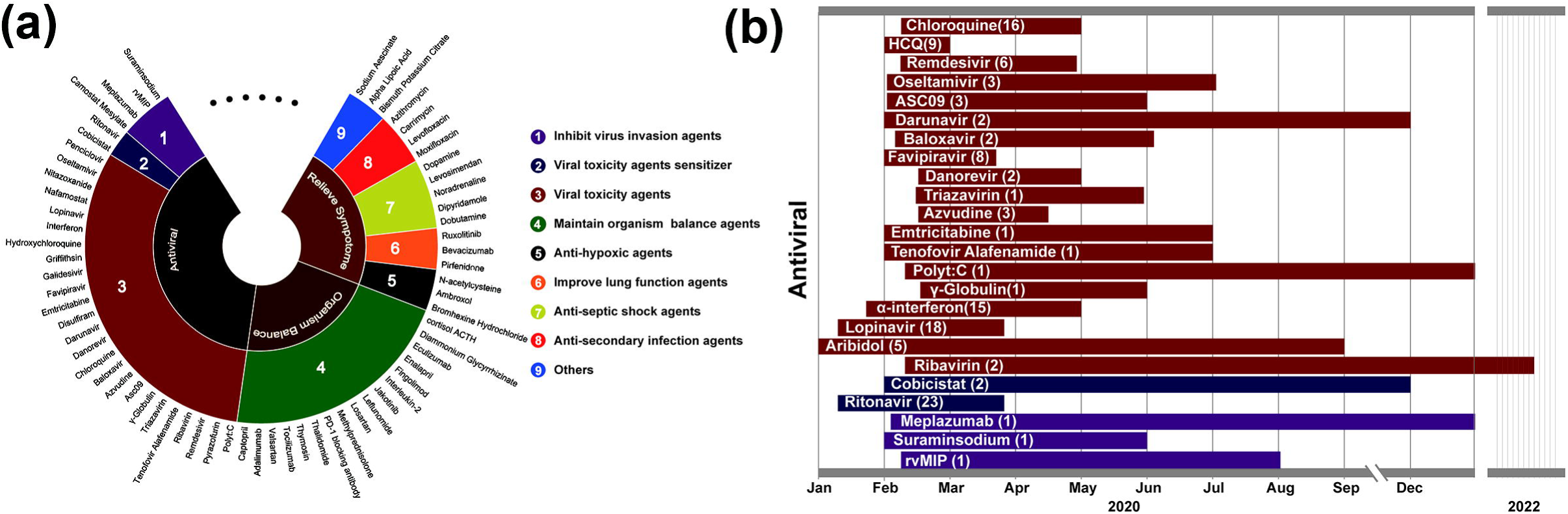
Candidate drugs for COVID-19 therapy. (a) All potential drugs which may be utilized for COVID-19 therapy were divided into three categories and nine sub-categories as indicated. (b) All clinical trials conducted for antivirus drugs were listed in the figure. The number in each column indicated the number of trials registered now (until Mar 3, 2020). The earliest initial date and the earliest endpoint which was expected by trial conductors of trials for a same drug were also indicted here.

### Drug-gene network for COVID-19 treatment

Genetic variation is one of the most powerful biomarkers to guide personalized therapy. Thus, it is important to systematically identify pharmacogenes which could affect response and toxicity of these 67 drugs. A drug-gene network which demonstrating the connection of each drug and pharmacogene was constructed (Figure 2A). After excluding drugs without pharmacogene data, a total of 44 drugs and 313 genes were connected.

**Figure 2:**
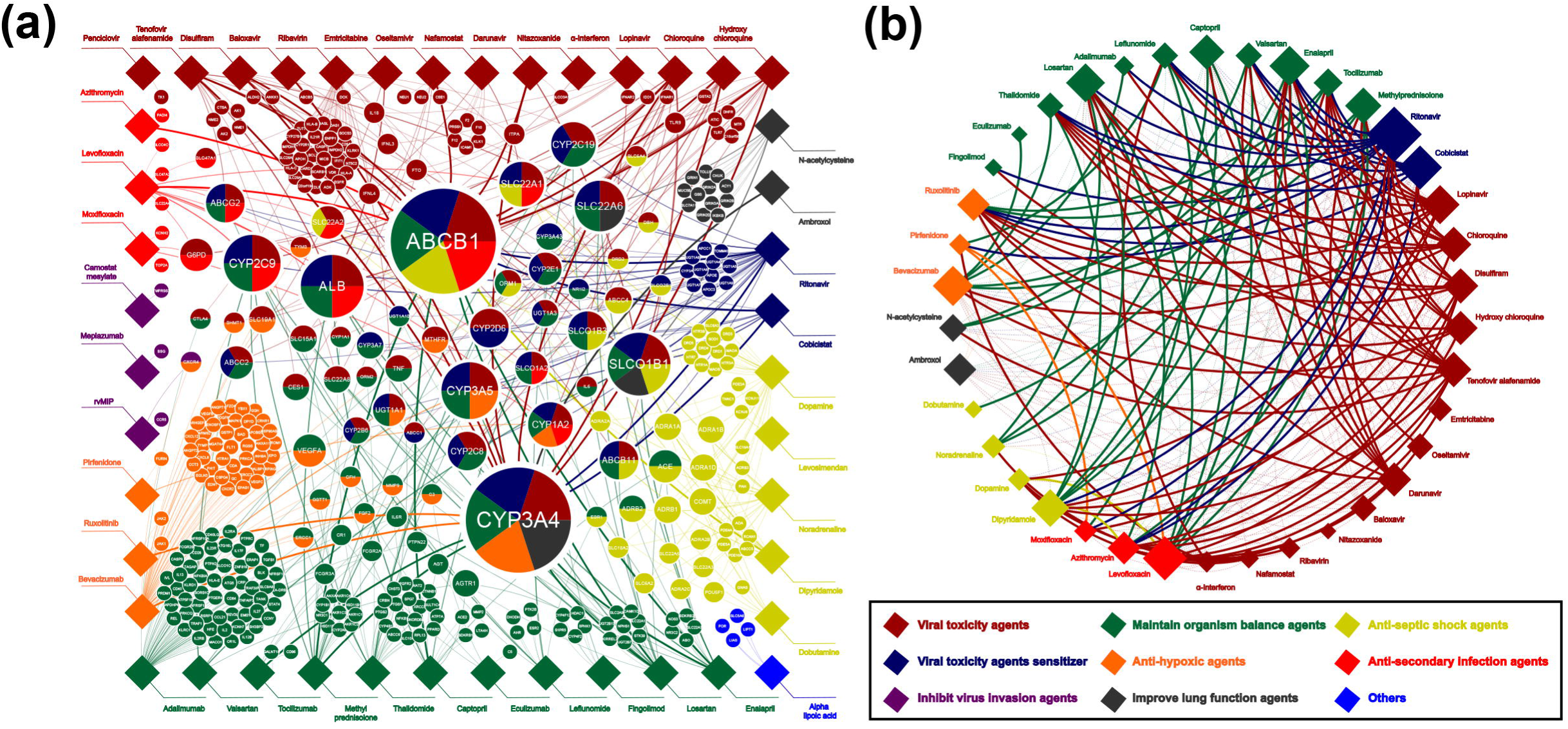
Drug-gene network in COVID-19 treatment. (a) The drug-gene network links 44 drugs and 313 candidate pharmacogenes as reported. All drugs were marked as diamond in the outermost, while all genes were marked as circles in the inner space. The colors of diamonds, circles and lines indicated the categories of related drugs in the drug-gene interactions. The kind of color of a circle reflects the number of drugs categories it links to. The size of the gene circle is determined by the number of linked drugs, and the transparency of the line it linked is proportional to the size of circles. (b) The DDIs predicted by drug-gene network. Solid lines indicate validated DDIs. The size of the diamond indicated the number of linked drugs. Drugs may interact with multi drugs should be used more carefully than other drugs.

Based on the nodes in this network, we could easily find genes shared by multiple drugs. They could be the mediator of drug-drug interactions (DDIs) (Figure 2B). In addition, their genetic variations could potentially affect response and toxicity of a number of different drugs. Thus, these genes should be paid attention during COVID-19 patient treatment. Among them, the top 2 genes both ligated 16 different drugs simultaneously. cytochrome P450 family (CYP) 3A4 is one of the major members of the cytochrome P450 enzymes superfamily, it involved in the metabolism of about half DCTs. The other one is ATP binding cassette subfamily B member 1 (ABCB1), which is an important transporter pumping out many drugs across the cellular membrane. Other top 10 genes included: solute carrier family solute carrier organic anion transporter family member 2B1, ALB, CYP3A5, CYP2C9, solute carrier family (SLC) 22A6, SLC22A1, CYP2C19 and CYP1A2. Most of them were related to absorption, distribution, metabolism and excretion. It is interesting to note that almost all of these gene are highly genetically variable and differ between populations

Another information obtained from this network was that there were actionable PGx biomarkers for three drugs based on CPIC or DPWG guidelines. They were G6PD and chloroquine hemolysis toxicity, vitamin D receptor (VDR) and ribavirin efficacy, inosine triphosphatase (ITPA) and ribavirin/α-interferon anemia risk, angiotensin I converting enzyme (ACE) and captopril response. We recommend that these genes should be preemptively tested before drug treatment.

### Mutation profiles of pharmacogenes

We next investigated the allelic frequency, location and function of genetic mutations for all pharmacogenes in different populations. A total number of 295,897 variations in these 313 genes were found in 125,748 subjects from GnomAD databases. Although the mutations distributed in all gene regions, the most common mutation types were intronic, non-synonymous and synonymous mutations (Figure 3A). It should be noteworthy that 32.8% of the mutations were non-synonymous, which is the major types of functional PGx variants. They could be the explanation of individual and ethnic difference for COVID-19 drug treatment. To learn the distribution of mutations in pharmacogenes in more detail. The fractions of different mutation types were provided for each gene. They were summarized based on the gene functions in Figure S2. The frequencies of these variations were indicated in Figure 3B, they showed similar distributions. However, there were remarkably differences for the total number of mutations.

**Figure 3:**
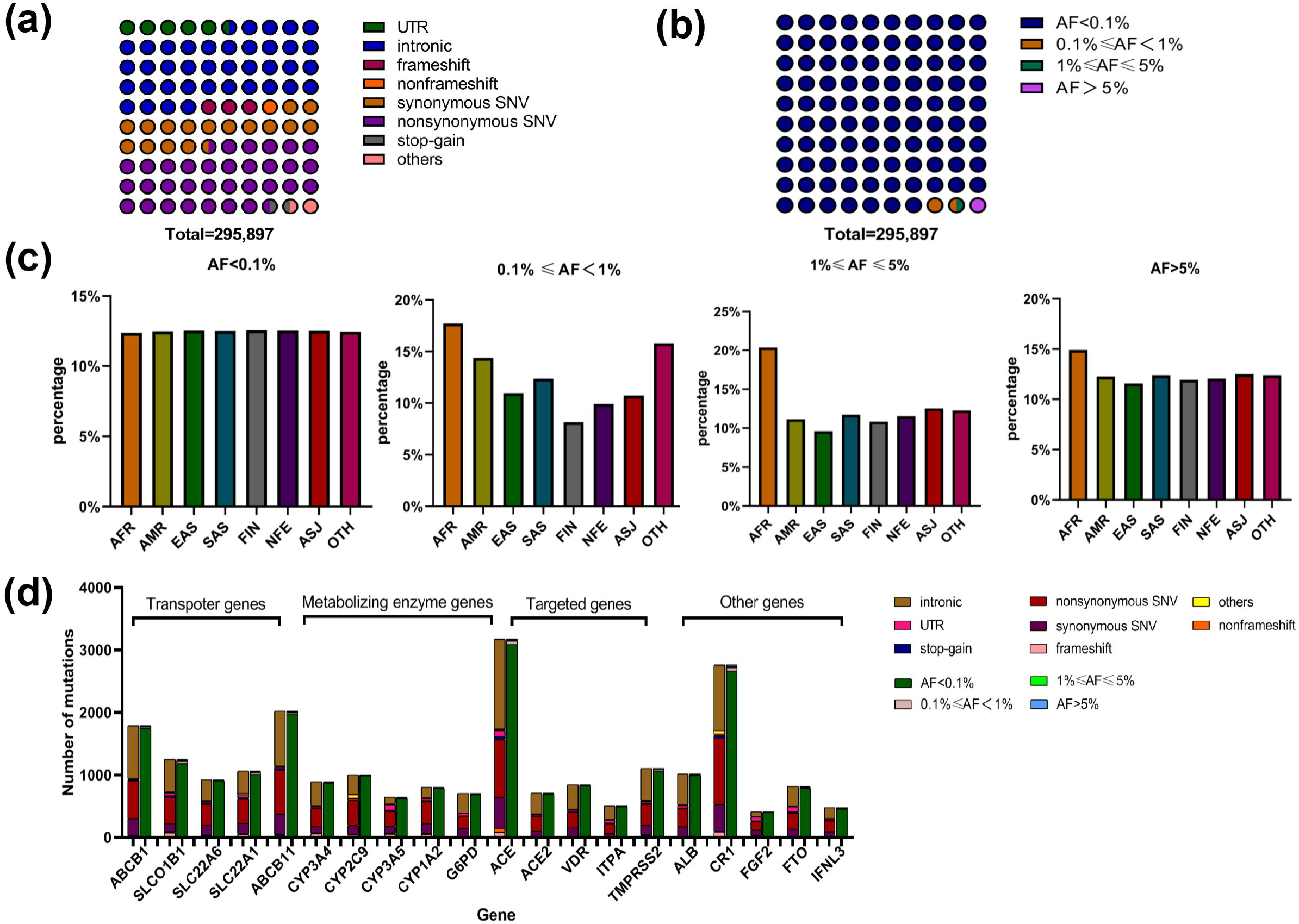
Variations profiles of pharmacogenes of drugs for COVID-19 therapy. (a)The distribution of all variations in pharmacogenes, which were classed according to their locations and functions. (b) The distribution of all variations in pharmacogenes, which were classed according to their frequency. (c) The proportion of variations with different frequency in eight different populations. (d) The number and distributions of variations in four different categories of important pharmacogenes.

Then, we explored the allelic frequencies of all mutations. As indicated in Figure 3B, 98.52% of the mutations were rare genetic variants with minor allele frequency (MAF) lower than 1%. This result emphasized again the importance of pharmacogenomic scale test before treatment. We further analyzed mutations with different MAF stratified by populations. Except African population, there was no significant difference of MAF distribution between the remained seven races (Figure 3C). The African individuals were higher genetically variable for these pharmacogenes, since they showed more percentages of common mutations (MAF>1%). This result indicated that they had higher requirement for PGx test.

Taken together, these results indicated that COVID-19 pharmacogenes were highly genetically variable, especially for African population. Thus, personalized drug treatment and PGx test is needed for different patients. In addition, these mutations distributed in all gene regions and most of them were rare variations (Figure 3D). Therefore, sequencing technologies is recommended for the preemptive test.

### Actionable and key PGx biomarkers

To provide clinical guide for COVID-19 treatment, we analyzed the actionable and key PGx biomarkers in detail. There are four paired actionable pharmacogenes and drugs: VDR and ribavirin, ITPA and ribavirin/α-interferon, G6PD and chloroquine, ACE and captopril.

Ribavirin is a classic antivirus drug which has been involved in the guideline for 2019-nCoV therapy as well. VDR polymorphism is associated with its efficacy (Figure 4A). There is only a common non-synonymous polymorphism M1T (rs2228570) that could hamper the VDR activity and the M1T carriers showed lower efficacy. The frequency of this mutation is over 0.5 in all populations (Figure 4A), which means more than half patients will be resistant to this drug.

**Figure 4:**
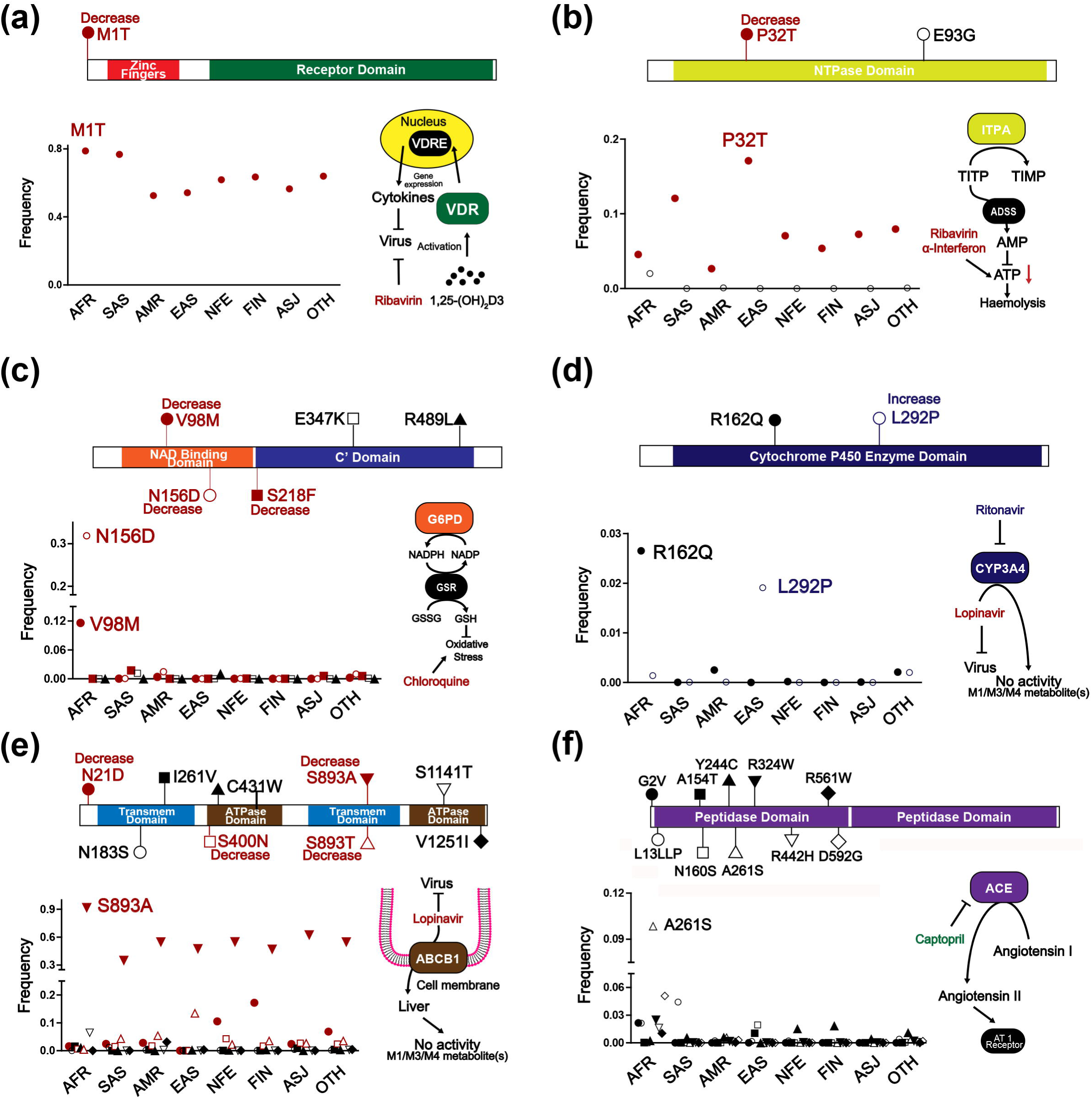
Common nonsynonymous variations in important pharmacogenes. Common nonsynonymous variations were indicated as amino alteration in the protein. The role of pharmacogenes in the treatment was indicated in the pathway. The frequencies of common nonsynonymous variations in different populations were also presented in the dot plot. Drug and pharmacogenes including: (a) VDR and ribavirin, (b)ITPA and ribavirin/α-interferon, (c) G6PD and chloroquine, (d) CYP3A4 and lopinavir, (e) ABCB1 and lopinavir, (f) ACE and captopril were presented here.

α-interferon is usually utilized in combination with antivirus drugs such as ribavirin in COVID-19 treatment. ITPA polymorphism was reported to be associated with the anemia risk in ribavirin/α-interferon treatment (Figure 4B). The functional mutation P32T (rs1127354) reduced the activity of the enzyme encoded by ITPA. The P32T carriers showed lower anemia risk. Another common non-synonymous polymorphism E93G (rs34982958) is also found in ITPA (Figure 4B), while its effect has not been revealed. The P32T was more identified in Asians (both of East Asians and South Asians, Figure 4B). Therefore, Asians may have a higher anemia risk in ribavirin/α-interferon treatment.

chloroquine has been utilized for 2019-nCoV therapy as guideline referred. G6PD polymorphisms has been reported to be associated with hemolysis risk (Figure 4C). There are two important variants V98M (rs1050828) and N156D (rs1050828), which could decrease the function of G6PD. The V98M and N156D carriers showed higher hemolysis risk for chloroquine treatment. In addition to these two mutations, we further identified three other common non-synonymous polymorphisms in all populations. Among them, the mutation S218F (rs5030868) was also reported to decrease the activity of G6PD, which indicated that this mutation could increase the hemolysis risk as well. What’s more, both V98M and N156D were more common in the African population (Figure 4C), suggesting that chloroquine is not recommended in African patients.

Lopinavir and ritonavir are used together to reduce their metabolism in liver through inhibiting the activity of CYP3A4, a very important enzyme which could metabolize about half of DCTs. The CYP3A4 polymorphism L292P (rs28371759, CYP*18B) was linked with increased CYP3A4 activity, thus facilitate the metabolism of drugs like lopinavir. L292P carriers may need to increase the dosage of these drugs compared with wild type patients (Figure 4D). Another common non-synonymous polymorphism R162Q in CYP3A4 was still not be reported with CYP3A4 phenotype now (Figure 4D). L292P was more in East Asians (Figure 4D), indicated that East Asians may metabolize lopinavir and ritonavir more rapidly.

As we mentioned above, ABCB1 can pumping out many drugs across the cellular membrane thus alleviate the efficacy. In our drug list, drugs like lopinavir can be transported out of cells by ABCB1, and its efficacy can be reduced (Figure 4E). We found there are 10 common non-synonymous polymorphisms in ABCB1 (Figure 4E). Among them, S893T, S893A (rs2032582), N21D (rs9282564) and S400N (rs2229109) were reported as deleterious mutations, which can increase the drug concentration through decreasing the efflux of ABCB1. S893A carriers were found in all populations with high frequency, which is up to 90% in Africans. S893T carriers were almost East Asians, while N21D carriers were Europeans (Figure 4E). These patients may be more response to these drugs transported by ABCB1.

In addition, ACE inhibitors (ACEI) can be used to improve the lung function in COVID-19 treatment. It was widely reported that ACE polymorphisms could affect ACEI therapeutic response (Figure 4F). The most important variant was I/D (rs1799752) polymorphism, which was a low frequency indel mutation causing low enzyme expression. The D allele carriers showed better response for ACEI treatment. In addition to this mutation, we further identified 10 common non-synonymous polymorphisms in all populations (Figure 4F). Although there is no related report about these variations, G2V (rs558593002), A154T (rs13306087), Y244C (rs3730025), R324W (rs35141294) and R561W (rs4314) were predicted to be loss-of-function mutations by PROVEAN. Thus, we speculated that patients harboring these single nucleotide polymorphisms (SNPs) were poor responders of this drug. Based on their MAFs in different populations, rs1799752 were more identified in the East Asian population (Figure 4F). Thus, we concluded that East Asian patients could be more resistance to this drug.

In summary, African patients could be more hazardous to chloroquine, Asian patients have a higher risk of anemia in ribavirin/α-interferon treatment and are more resistant to captopril. Meanwhile, drugs like lopinavir and ritonavir which metabolized by CYP3A4 or transported by ABCB1 should be carefully utilized in East Asians.

### COVID-19 treatment based on sc-RNA seq data

Recently, sc-RNA seq technology was rapidly developed. It is capable to specifically profile cell populations at the single-cell resolution. Thus, it transformed many fields of genomic research. Here, we tried to explore the utility of sc-RNA seq on personalized COVID-19 treatment. We collected healthy adults sc-RNA seq data from 4 lung and 5 liver tissues.

Lung is the major 2019-nCoV attacking organ. Thus, we analyzed the expression of virus or drug target genes in different lung cells (Figure 5A). Based on the current findings, ACE2 and transmembrane serine protease 2 (TMPRSS2) were the two major targets of both virus and some drugs. As indicated in Figure 5B, TMPRSS2 was almost expressed in all kinds of cells. In our enrolled drugs, camostat mesylate act as a trypsin like protease inhibitor and could be potentially used to anti-virus by inhibiting TMPRSS2 activity. Due to the ubiquitous expression of its target, camostat mesylate could also cause potential toxicity when exerting therapeutic effect. On the other hand, ACE2 was mainly expressed in type I pneumocyte, type II pneumocyte, FOXN4^+^, club and mast cells. chloroquine phosphate can change the ACE2 structure, or inhibit the binding of coronavirus S protein with ACE2. Based on these data, chloroquine was more effectively targeting drugs with less side effects (Figure 5C). Sc-RNA seq data in lung tissue suggested that ACE2 targeting drugs (chloroquine) were superior for COVID-19 treatment than TMPRSS2 targeting drugs (camostat mesylate).

**Figure 5:**
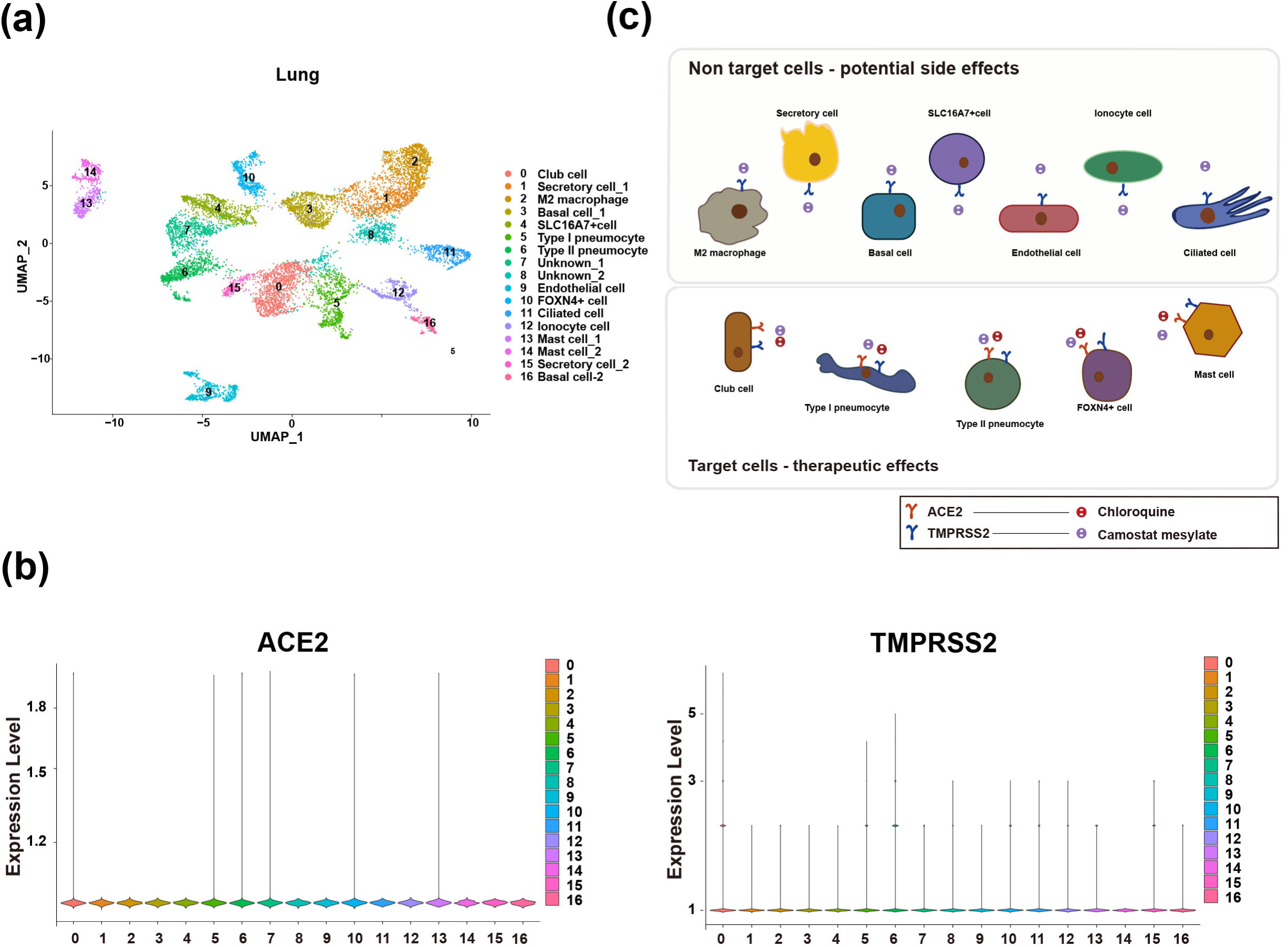
Discrepancy of pharmacogenes’ expression in target genes. (a) Lung cells were clustered by UMAP method. Different cell populations were marked in different colors. (b) Violin plot of ACE2 and TMPRSS2 expression in different cell types of lungs. (c) Mechanism of chloroquine and Camostat mesylate acting on different lung cells by identifying targets.

Most drugs can be metabolized in the liver. We next analyzed the expression of drug metabolic enzymes in different liver cells (Figure 6A). CYP1A2, CYP2E1, CYP2C8, CYP2C9, CYP2B6, CYP2D6, CYP3A4 and CYP3A5 were the major CYP450 enzymes responsible for 67 drugs metabolism. Their expression was investigated (Figure 6B, 6D). In addition to hepatocytes, it was unexpected to found that some enzymes were also found to be highly expressed in immune cells. Based on the expression distribution, we categorized all enzymes into two groups: (1) narrow distribution enzymes (NDE) which were only expressed in hepatocytes and scarcely expressed in other cells, including CYP3A4, CYP1A2, CYP2B6, CYP4F2 and CYP4F12; (2) wide distribution enzymes (WDE) which is expressed in both hepatocytes and immune cells in the liver, including CYP2C8, CYP2C9, CYP2D6 and CYP3A5 (Table 1). For our enrolled drugs, anti-viral agents such as α-interferon, ambroxol, baloxavir and darunavir were mainly metabolize by NDEs, while thalidomide and valsartan were mainly metabolized by WDEs, and some drugs such as chloroquine and leflunomide were metabolize by both NDEs and WDEs. Currently, the consequence of drug metabolism in immune cells still remains unknown. However, it was rational to speculate that the toxically drug metabolites in the immune cells would affect liver immunologic function. Thus, in case of producing toxically metabolites, the drugs that can be metabolized by WDE would be more tending to cause hepatotoxicity. They should be cautiously used when alternative drugs were available. Leflunomide can be metabolized by WDEs CYP2C9 which expressed both in hepatocyte and CD4 ^+^ T cells. Its active metabolite, A771726, inhibits cell survival and proliferation by inhibiting pyrimidine synthesis. The aggregation of A771726 in CD4 ^+^ T cells inhibits cell survival and proliferation, and damages the liver immunologic function (Figure 6C). Therefore, leflunomide may be not recommended while other immunosuppressive drugs are available.

**Table 1:**
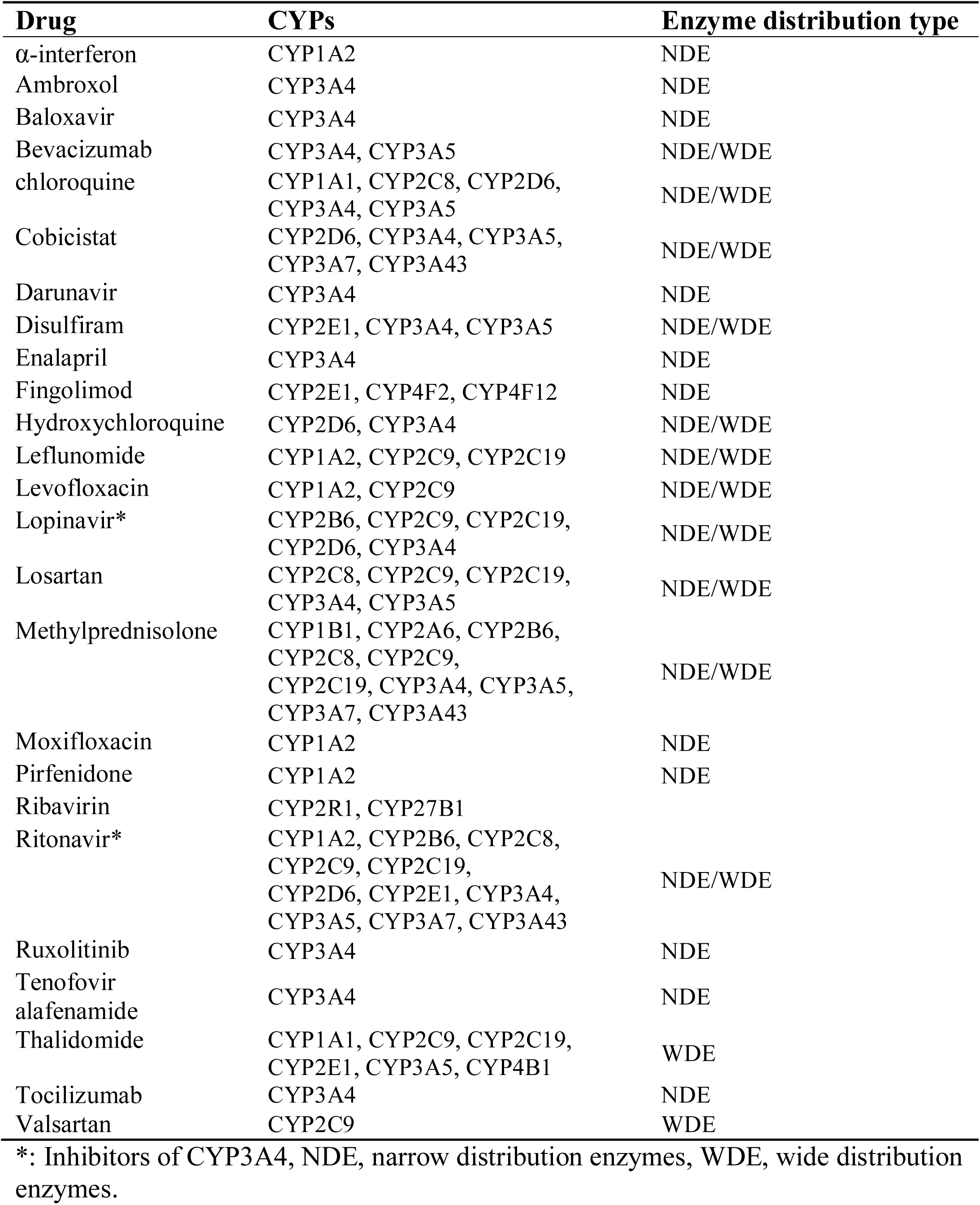
CYP450s profiles in COVID-19 treatment.

**Figure 6:**
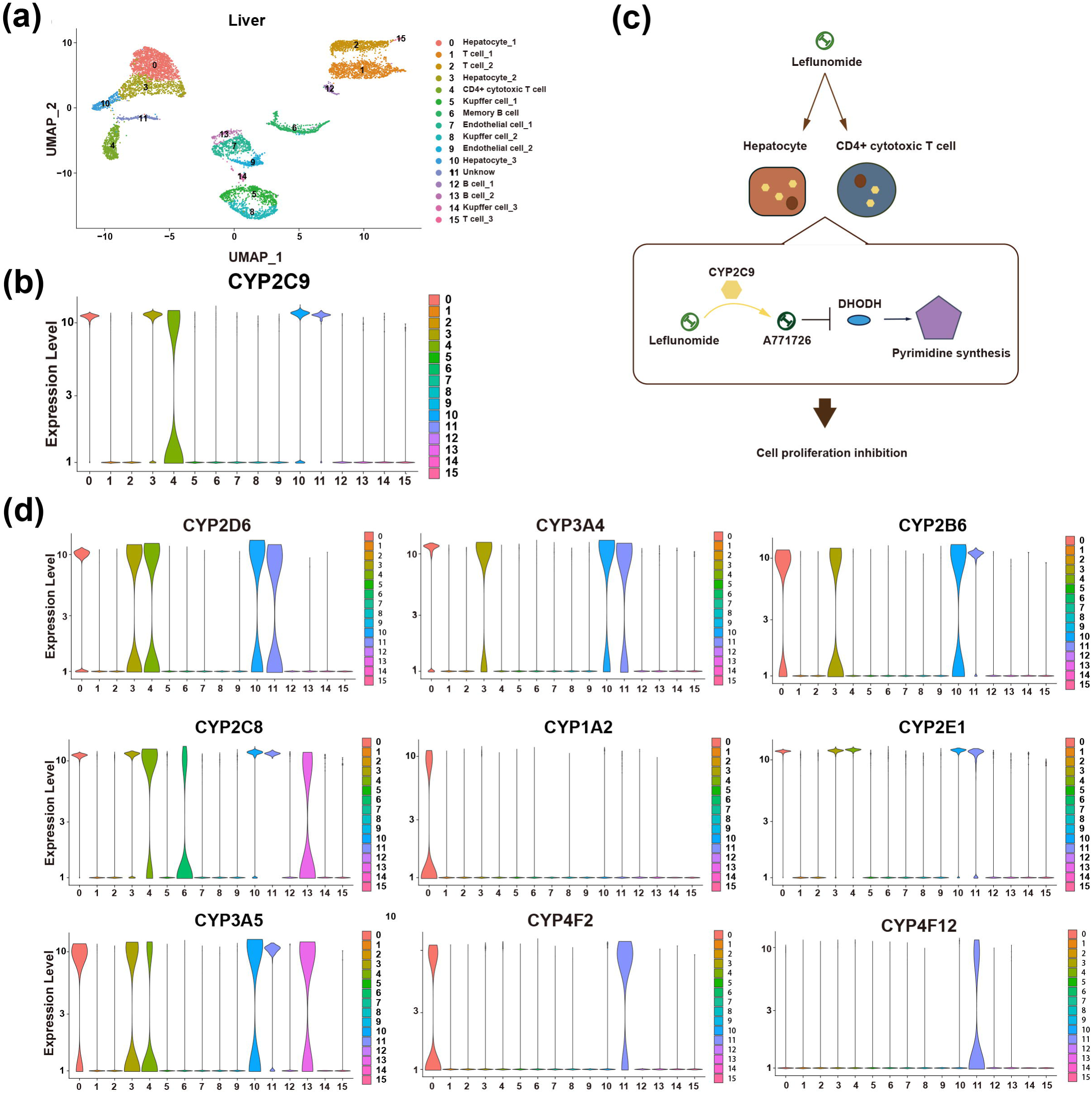
Discrepancy of pharmacogenes’ expression in CYP450 enzymes. (a) Liver cells were clustered by UMAP method. (b) Violin plot of CYP2C9 expression in different cell types of livers. (c) The mechanism of Leflunomide inhibiting cell proliferation after metabolism by CYP2C9. (d) Violin plot of CYP2D6, CYP3A4, CYP2B6, CYP2C8, CYP1A2, CYP2E1, CYP3A5, CYP4F2 and CYP4F12 expression in different cell types of livers.

### Candidate Strategies for COVID-19 Therapy

Since there are many candidate drugs for COVID-19 therapy, it is important to make a better choice for each patient. For example, ribavirin and α-interferon are not suggested in patients with M1T variation in VDR or without P32T variation in ITPA, then other anti-virus drugs can be used in these patients. Similarly, chloroquine is not suggested to be utilized in patients with V98M and N156D variants. To avoid these abuses of drugs in COVID-19 therapy, a genetic detection panel is suggested. In this panel, SNPs with clinic evidences in pharmacogenes should be considered. The mutation frequencies are not the same in all populations, and we provided the frequency of candidate pharmacogenes SNPs in all populations (Figure 7). Mutations like rs117648444 is only found in FIN, so that the panels for FIN should include this variation. Meanwhile, suggestions should be given based on these SNPs. Examples can be found in Table 2.

**Table 2:**
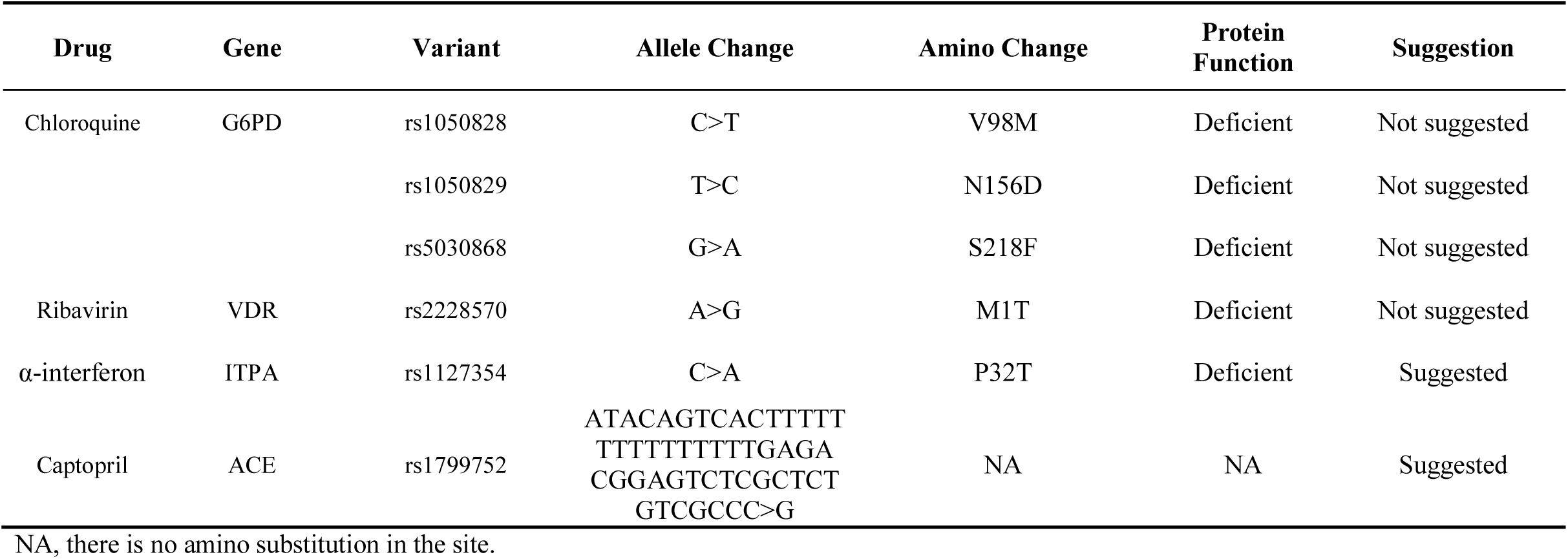
Suggestions for precision medicine based on nonsynonymous variations in COVID-19 treatment.

**Figure 7:**
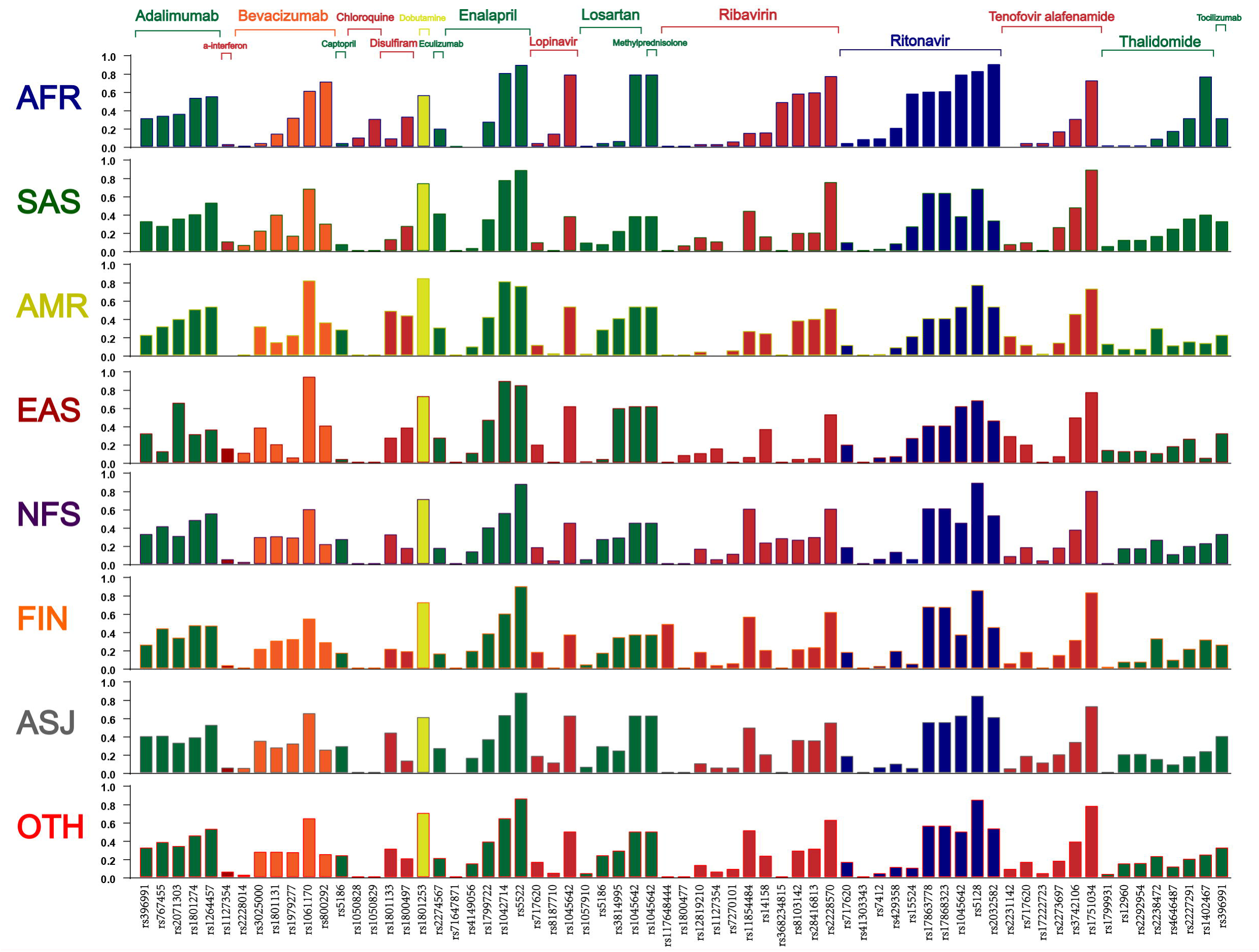
The frequency of important SNPs in eight populations. Drugs were listed in the top of the panel, while the SNPs were listed in the bottom of the panel. The color of the column is as same as the color of the drug, which indicated the category of the drug.

As a large part of drugs are metabolized by CYP450 in liver, the activity of CYP450 family may impact many drugs metabolization rate. Due to the activity difference exists among populations, the genotypes of important alleles in CYP450 family should be detected after diagnosis, then treatment strategies for these drugs can be suggested in these patients. Meanwhile, DDI should also be considered as many drugs may be utilized together during COVID-19 treatment as mentioned above. For instance, drugs like lopinavir and ritonavir can inhibit the activity of CYP3A4, thus the drugs dosage should be adjusted when combined with drugs metabolized by CYP3A4. Drugs that were metabolized by CYP450 family in COVID-19 therapy were listed in Table 3. In addition, when chloroquine is utilized, drugs like losartan which can increase the concentration of chloroquine may be replaced by valsartan. Validated DDIs between drugs for COVID-19 treatment are given in table 3.

**Table 3:**
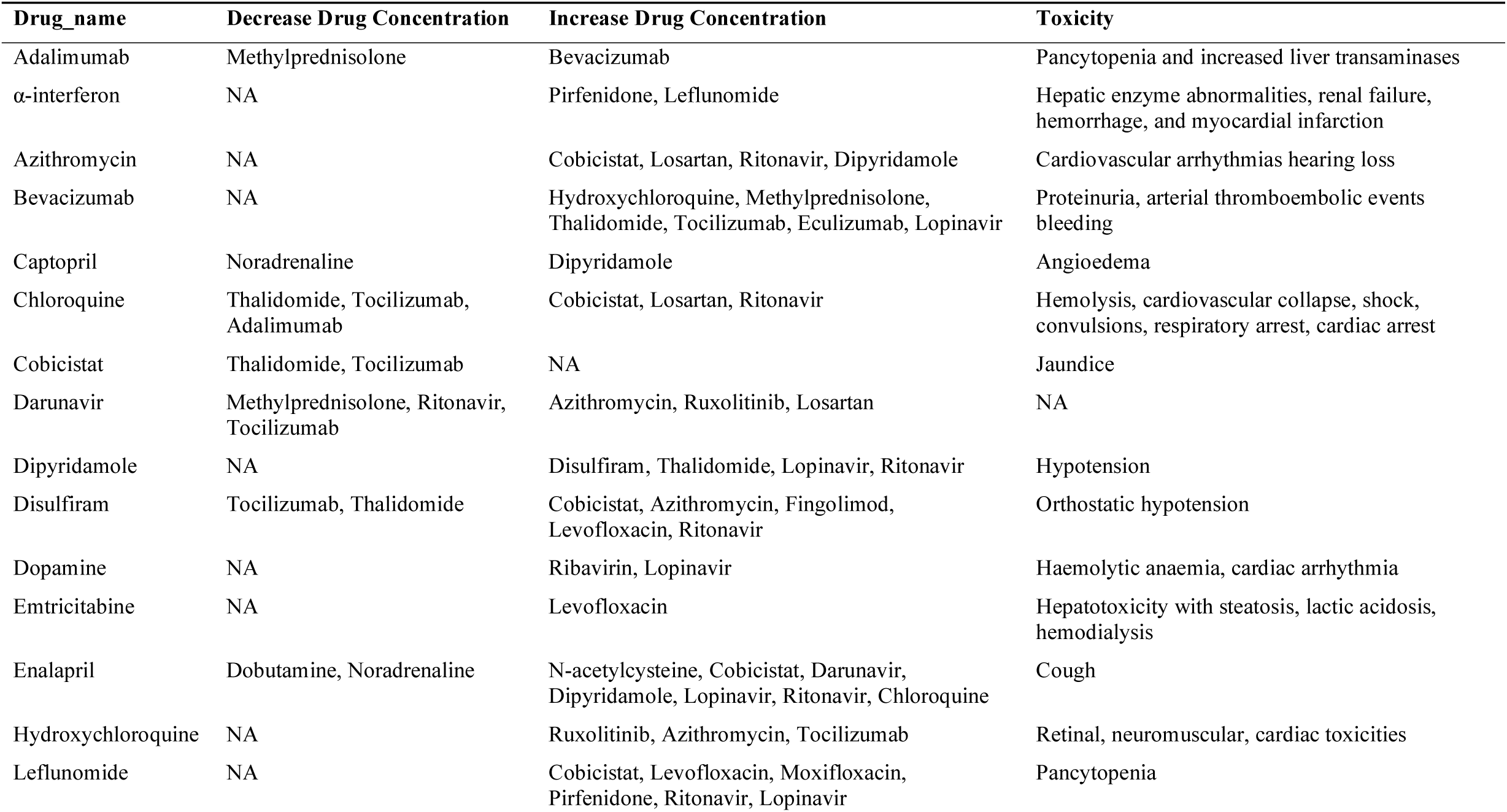
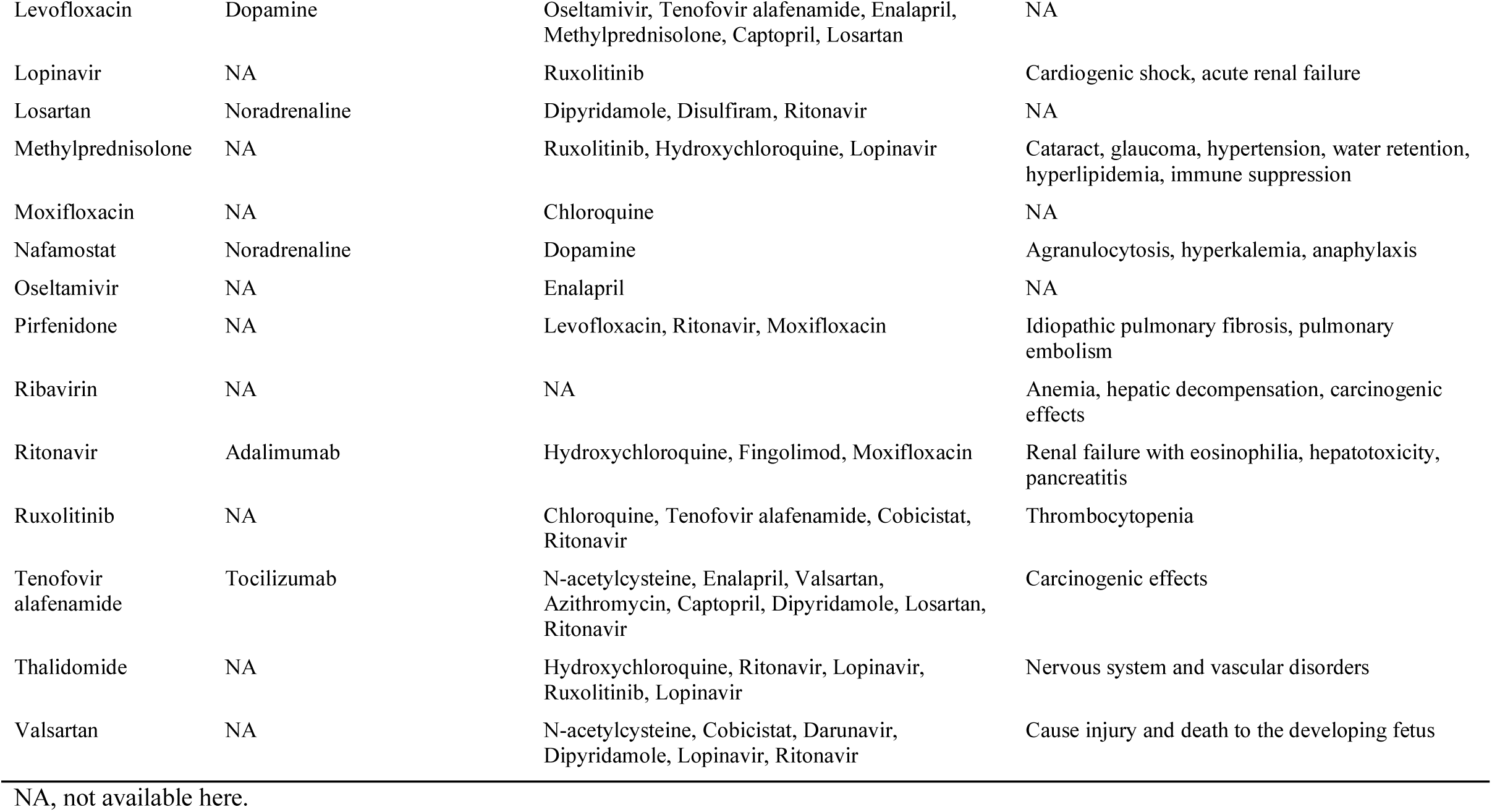
Validated DDIs and toxicity.

| **Drug_name** | **Decrease Drug Concentration** | **Increase Drug Concentration** | **Toxicity** |
| --- | --- | --- | --- |
| Adalimumab | Methylprednisolone | Bevacizumab | Pancytopenia and increased liver transaminases |
| α-interferon | NA | Pirfenidone, Leflunomide | Hepatic enzyme abnormalities, renal failure, hemorrhage, and myocardial infarction |
| Azithromycin | NA | Cobicistat, Losartan, Ritonavir, Dipyridamole | Cardiovascular arrhythmias hearing loss |
| Bevacizumab | NA | Hydroxychloroquine, Methylprednisolone, Thalidomide, Tocilizumab, Eculizumab, Lopinavir | Proteinuria, arterial thromboembolic events bleeding |
| Captopril | Noradrenaline | Dipyridamole | Angioedema |
| Chloroquine | Thalidomide, Tocilizumab, Adalimumab | Cobicistat, Losartan, Ritonavir | Hemolysis, cardiovascular collapse, shock, convulsions, respiratory arrest, cardiac arrest |
| Cobicistat | Thalidomide, Tocilizumab | NA | Jaundice |
| Darunavir | Methylprednisolone, Ritonavir, Tocilizumab | Azithromycin, Ruxolitinib, Losartan | NA |
| Dipyridamole | NA | Disulfiram, Thalidomide, Lopinavir, Ritonavir | Hypotension |
| Disulfiram | Tocilizumab, Thalidomide | Cobicistat, Azithromycin, Fingolimod, Levofloxacin, Ritonavir | Orthostatic hypotension |
| Dopamine | NA | Ribavirin, Lopinavir | Haemolytic anaemia, cardiac arrhythmia |
| Emtricitabine | NA | Levofloxacin | Hepatotoxicity with steatosis, lactic acidosis, hemodialysis |
| Enalapril | Dobutamine, Noradrenaline | N-acetylcysteine, Cobicistat, Darunavir, Dipyridamole, Lopinavir, Ritonavir, Chloroquine | Cough |
| Hydroxychloroquine | NA | Ruxolitinib, Azithromycin, Tocilizumab | Retinal, neuromuscular, cardiac toxicities |
| Leflunomide | NA | Cobicistat, Levofloxacin, Moxifloxacin, Pirfenidone, Ritonavir, Lopinavir | Pancytopenia |
| Levofloxacin | Dopamine | Oseltamivir, Tenofovir alafenamide, Enalapril, Methylprednisolone, Captopril, Losartan | NA |
| Lopinavir | NA | Ruxolitinib | Cardiogenic shock, acute renal failure |
| Losartan | Noradrenaline | Dipyridamole, Disulfiram, Ritonavir | NA |
| Methylprednisolone | NA | Ruxolitinib, Hydroxychloroquine, Lopinavir | Cataract, glaucoma, hypertension, water retention, hyperlipidemia, immune suppression |
| Moxifloxacin | NA | Chloroquine | NA |
| Nafamostat | Noradrenaline | Dopamine | Agranulocytosis, hyperkalemia, anaphylaxis |
| Oseltamivir | NA | Enalapril | NA |
| Pirfenidone | NA | Levofloxacin, Ritonavir, Moxifloxacin | Idiopathic pulmonary fibrosis, pulmonary embolism |
| Ribavirin | NA | NA | Anemia, hepatic decompensation, carcinogenic effects |
| Ritonavir | Adalimumab | Hydroxychloroquine, Fingolimod, Moxifloxacin | Renal failure with eosinophilia, hepatotoxicity, pancreatitis |
| Ruxolitinib | NA | Chloroquine, Tenofovir alafenamide, Cobicistat, Ritonavir | Thrombocytopenia |
| Tenofovir alafenamide | Tocilizumab | N-acetylcysteine, Enalapril, Valsartan, Azithromycin, Captopril, Dipyridamole, Losartan, Ritonavir | Carcinogenic effects |
| Thalidomide | NA | Hydroxychloroquine, Ritonavir, Lopinavir, Ruxolitinib, Lopinavir | Nervous system and vascular disorders |
| Valsartan | NA | N-acetylcysteine, Cobicistat, Darunavir, Dipyridamole, Lopinavir, Ritonavir | Cause injury and death to the developing fetus |

NA, not available here.

Finally, based on current researches, 2019-nCoV can only infect cells with ACE2. As indicated above, ACE2 is not expressed in all cells, while TMPRSS2 can be found in large part of cells. So that drugs like camostat mesylate is not recommended compared to ACE2 specific inhibitors due to the side effect caused by TMPRSS2 in uninfected cells.

## Discussion

This study firstly focused pharmacogenetics and provided clinical suggestions for precision medicine in COVID-19 treatment systematically. 44 DCTs were showed to associated with at least one gene. And drug-gene network highlighted multi drug related genes in COVID-19 treatment. Meanwhile, inter-racial variances in drug efficacy were found related to nonsynonymous mutations among different races, which indicated that racial special strategy for COVID-19 therapy should be considered during the outbreak of COVID-19 worldwide. Since nonsynonymous mutations and gene expression levels can both affect the DCTs efficacy and toxicity as mentioned above, optimized therapy strategies are needed to benefit these affected patients all over the world. In this study, we give three suggestions for DCTs as follows. Firstly, drugs in CPIC guideline including ribavirin, α-interferon, chloroquine and captopril should be utilized with genetic detections as guided. In this situation, adequate gene detection kits should be prepared in Africa for COVID-19 therapy, as related genetic variation frequency is higher in AFRs than other countries. Secondly, drug such as chloroquine whose efficacy and toxicity can be determined by nonsynonymous mutations, may be preferred in special population with lower frequency of risk alleles such as Finnish and Non-Finnish European. Otherwise, alternative drug for chloroquine can be used in high risk populations. Thirdly, for drugs that can inhibit the activity of CYP450, DDI should be prevented in COVID-19 treatment (Table 1). When inhibitors of CYP450 were utilized, the dosage of these drugs should be adjudged carefully to avoid overdosage.

Although precision medicine should be considered in COVID-19 treatment, there are still many challenges to face. The first challenge is that pharmacogenes of many drugs for COVID-19 are still not clear, due to that many newly developed drugs are still under clinical trials, their pharmacogenes can hardly be fully identified currently. A potential solution is to predict candidate pharmacogenes according to the chemical structure of a new drug by the artificial intelligence and machine learning algorithm ^[21, 22]^. Then related molecular biological studies can be conducted to validate these candidate pharmacogenes rapidly. Then the period of pharmacogenes identification can be shorted. The second challenge is that some serious nonsynonymous mutations can be hardly identified in current pharmacogenetics studies, due to the frequencies of many nonsynonymous mutations are lower than 1%. That’s why the nonsynonymous mutations we presented above are common variants. If there are only a few patients carried a serious nonsynonymous mutation, related study can hardly be conducted to identify this mutation because of the limited sample size. However, as we indicated in Figure 3, most of nonsynonymous mutations are rare variations. Therefore, a reliable way to predict potential pharmacogenetic variation is also important to improve the efficacy or avoid serious toxicity ^[23]^. The third challenge is that a suitable strategy for precision medicine in COVID-19 treatment is still lack. A well-designed gene detection panel is important to fully characterized the patients’ genotypes which relate to the drug effects ^[24]^. Then the best treatment regimen can be utilized for this patient, and both of the efficacy and the safety can be guaranteed. In our study, feasible suggestions were given, and it still can be much improved with the development of some pharmacogenetic studies in the future. Last but not least, the expression level of pharmacogenes in patients can hardly be detected in single cell resolution during COVID-19 treatment, although the expression level of these genes can be associated with drug efficacy or toxicity. A potential solution is to build a systematic pharmacogenes expression map in single cell level among different populations in different age stages ^[25]^. This map would be helpful for personalized medicine in the future.

## Data Availability

All data are presented in supplementary data.

## Acknowledgment

This work was supported by the National Natural Science Foundation of China (81773823, 81573463, 81974511), National Science and Technology Major Project of China (2017ZX09304014, 2019ZX09201-002-006).

## Abbreviations

ARDS: acute respiratory distress syndrome
ABCB1: ATP binding cassette subfamily B member 1
ACE: angiotensin I converting enzyme
ACEI: ACE inhibitors
COVID-19: coronavirus disease 2019
CYP: cytochrome P450 family
CPIC: Clinical Pharmacogenetics Implementation Consortium
DCTs: drugs for COVID-19 therapy
DDIs: drug-drug interactions
G6PD: glucose-6-phosphate dehydrogenase.
ITPA: inosine triphosphatase
NDE: narrow distribution enzymes
PGx: pharmacogenetics
SNP: single nucleotide polymorphism
SLC: solute carrier family
SLCO: SLC organic anion transporter family
Sc-RNA seq: single cell RNA-sequencing
TMPRSS2: transmembrane serine protease 2
VDR: vitamin D receptor
WDE: wide distribution enzymes
2019-nCoV: 2019 novel coronavirus

